# COVID-19 vaccine coverage among immigrants and refugees in Alberta: a population-based cross-sectional study

**DOI:** 10.1101/2022.04.11.22273644

**Authors:** Shannon E. MacDonald, Yuba Raj Paudel, Crystal Du

**Author notes:** **Address correspondence to:** Shannon MacDonald, Faculty of Nursing, Level 3, Edmonton Clinic Health Academy, 11405-87 Ave, Edmonton, Alberta, Canada, T6G 1C9, [ ].

## Abstract

**Introduction:** Studies have shown that immigrants have lower vaccination rates than the Canadian-born population. We sought to assess COVID-19 vaccine coverage and factors associated with uptake among foreign-born immigrants relative to the non-immigrant population in Alberta, Canada.

**Methods:** In this cross-sectional study, we analyzed population-based linked administrative health data from Alberta to examine vaccine coverage for 3,931,698 Albertans, of which 731,217 were immigrants. We calculated COVID-19 vaccination coverage as the proportion of eligible Albertans with a record of receiving at least one dose of a COVID-19 vaccine as of November 29, 2021. We used multivariable logistic regression to examine the association of vaccine coverage with migration status (immigrants: four categories based on time since migration and non-immigrants) adjusting for socio-demographic variables.

**Results:** Overall, COVID-19 vaccination coverage was higher among immigrants (78.2%; 95% CI: 78.1%-78.3%) compared to non-immigrants (76.0%; 95% CI: 75.9%-76.0%). Coverage among immigrants differed by continent of origin, with North America, Oceania, and Europe having the lowest coverage. Although vaccine coverage was relatively uniform across neighborhood income quintiles for immigrants, immigrants living in rural areas had lower vaccine coverage compared to non-immigrants living in rural areas. Multivariable logistic regression analysis showed a significant interaction between age category and migration status. While immigrants below 50 years of age generally had significantly higher vaccine coverage compared to non-immigrants, there was some variation based on time since migration. Immigrants above 50 years of age showed significantly lower coverage compared to non-immigrants of the same age.

**Conclusion:** Public health interventions should focus on older immigrants, immigrants living in rural areas, and immigrants from specific continental backgrounds in order to improve COVID-19 vaccination coverage.

## Introduction

Control of the COVID-19 pandemic in Canada is dependent on most of the population receiving COVID-19 vaccination.(1) More than one in five (21.9%) people in Canada are foreign-born immigrants.(2) Exposure on the front lines, and working and living in densely populated areas, have contributed to higher COVID-19 infections among immigrants.(3) Thus, it is critical to ensure high COVID-19 vaccination coverage in immigrant populations.

Studies have shown that immigrant populations in Canada tend to have lower vaccination rates for some routine vaccines and higher vaccine-preventable disease-related hospitalizations than the Canadian-born population.(4,5) Specific barriers to vaccination for immigrants include cultural factors, knowledge barriers, inadequate health care access, and vaccine hesitancy; barriers specific to COVID-19 vaccination include language barriers and novelty of the vaccines.(6,7)

In the province of Alberta, as in most of Canada, COVID-19 vaccines have been available free-of-charge, irrespective of immigration status. Roll-out has occurred in a phased manner, based on occupational risk and age.(8) COVID-19 vaccines are provided by public health nurses, pharmacists, physicians, and other health care providers. COVID-19 vaccine coverage in Alberta varies by age and place of residence,(9) but there is limited information on coverage in the immigrant population compared with the non-immigrant population.

Given the large size of the immigrant population and their potentially higher risk for COVID-19 infection, it is imperative to assess COVID-19 vaccination coverage for this population. It is also important to understand factors associated with vaccine uptake, so that appropriate and timely actions can be taken to improve coverage. Thus, the primary objectives of this study were to [1] compare COVID-19 vaccine coverage between foreign-born immigrants and refugees in Alberta to Canadian-born residents, and [2] to measure the association of time since migration with uptake of at least one dose of a COVID-19 vaccine.

## Methods

### Setting

Alberta is a western Canadian province of 4.5 million residents, 99% of whom are registered with the publicly funded Alberta Health Care Insurance Plan (AHCIP). While the Canadian government oversees vaccine approval and procurement, provinces are responsible for the roll-out of vaccination programs.(10,11) COVID-19 vaccination programs in Alberta began in December 2020 in a phased manner, with vaccine availability to the entire population aged 12 and above by mid-May 2021. Records for all COVID-19 vaccines administered in the province, regardless of provider, are submitted to the provincial immunization repository, known as Imm/ARI (with a few exceptions, noted below).

### Study design, population, and data sources

This was a population-based cross-sectional study using administrative data held by the Alberta Ministry of Health and including all residents of Alberta aged 12 years and above. Albertans <12 years were excluded because they were not eligible to receive the COVID-19 vaccine during the study period. We used the AHCIP quarterly population registry for the last quarter of 2020 to identify residents of the province, irrespective of birthplace. We excluded First Nations residents of Alberta (since data was not consistently submitted to Imm/ARI), Lloydminster residents (since vaccines are delivered by the neighbouring province), and those who left the province or died during the study period. Figure A1 in the appendix provides further detail on exclusions. We used the provincial immigrant registry to identify foreign-born immigrants and refugees who migrated to Alberta from another country between 1983 and 2020. We considered interprovincial migrants as non-immigrants since it is impossible to differentiate between foreign-born immigrants and Canadian-born individuals moving to Alberta from another province. We used the Imm/ARI repository to determine COVID-19 vaccination status. We extracted data on November 29, 2021, then deterministically linked the databases using unique personal health numbers.

### Outcome measure

We defined ‘COVID-19 vaccination coverage’ as the proportion of eligible Alberta residents who received at least one dose of a COVID-19 vaccine and ‘full COVID-19 vaccination coverage’ as the proportion who received at least two doses.

### Exposure variables

We created a migration status variable with five categories, including non-immigrants and four categories of immigrants based on year since migration: (1) in or after Jan 2019 (i.e., in the past 2 years), (2) Jan 2011 to Dec 2018 (i.e., in the past 3-10 years), (3) Jan 2000 to Dec 2010 (i.e., in the past 11-20 years), (4) Jan 1983 to Dec 1999 (i.e., more than 20 years ago).

We grouped participants into six age categories based on the categories used to prioritize COVID-19 vaccine eligibility in Alberta: 12-17 years, 18-29 years, 30-49 years, 50-64 years, 65-74 years, and 75 years and above. Biologic sex at birth was categorized into male and female. Neighborhood income quintiles (Q1 indicating the poorest neighborhood and Q5 indicating the richest) were based on the 2016 Canadian census. Place of residence was categorized into three categories based on the 2016 census: metro and moderate metro, urban and moderate urban, and rural and remote rural. Place of origin among the immigrant population was divided into six continental categories: North America, South America, Europe, Asia, Africa, and Oceania.

## Statistical analysis

We measured vaccination coverage among immigrants by continent of origin and age category. We also stratified coverage based on migration status (time since migration or non-immigrant) and compared vaccination coverage by age category, sex, place of residence, and income quintile. Those with missing data on key sociodemographic variables (age, gender, and postal code) were excluded from analysis. We used multivariable logistic regression to adjust for possible confounders associated with the outcome (uptake of at least one dose of a COVID-19 vaccine). Variables adjusted for in the multivariable model included: age category, sex, place of residence, and income quintile. Before running the multivariable model, we tested for multicollinearity among exposure variables and for plausible interactions. Since a strong interaction was detected between the age category and migration status, interaction terms for migration status and age category were included in the final model.

We performed a sensitivity analysis using full vaccination coverage (i.e., two or more doses) as the outcome in the multivariable model. For this sensitivity analysis, we excluded those who received the first dose of a COVID-19 vaccine after October 15, 2021 to allow everyone in the cohort to have at least the minimum required interval (42 days) between two doses.

We performed statistical analysis using SAS 9.4 (SAS Institute Inc., Cary, NC) with statistical significance set at p<0.05. This research, involving data from human participants, conformed to the principles embodied in the Declaration of Helsinki and was granted ethical approval by the University of Alberta Health Research Ethics Board (Ethics ID: Pro00114786).

### Patient and public involvement

Patients and the public were not involved in the design, conduct and dissemination of this research.

## Results

### Cohort characteristics relative to migration status

After excluding participants with missing data on time since migration (n=13), postal codes (n=117), and sex (n=8) from age-eligible Albertans, the final sample size was 3,931,698, of which 731,217 (18.5%) were international immigrants to Alberta. Those above 50 years of age comprised nearly 30% of the immigrant population and 40% among non-immigrants. Females comprised 51.8% of immigrants and 49.2% of non-immigrants. Immigrants from Asia comprised nearly half of the immigrant population in this study (48.3%).

### COVID-19 vaccine coverage

Overall, COVID-19 vaccine coverage was slightly higher among immigrants (78.2%; 95% CI: 78.1%, 78.3%) compared to non-immigrants (76.0%; 95% CI:75.9%, 76.0%). Coverage among immigrants differed by continent of origin (Table 1). Asian immigrants had the highest overall vaccine coverage (83.3%) followed by African immigrants (79.3%). In contrast, immigrants from elsewhere in North America had the lowest overall coverage (62.9%). For the age categories between 12 and 74 years, immigrants from North America, Oceania, and Europe had the lowest coverage. For the oldest age group (75 years and older), immigrants from North America, Asia, and Europe had the lowest coverage.

**Table 1.**
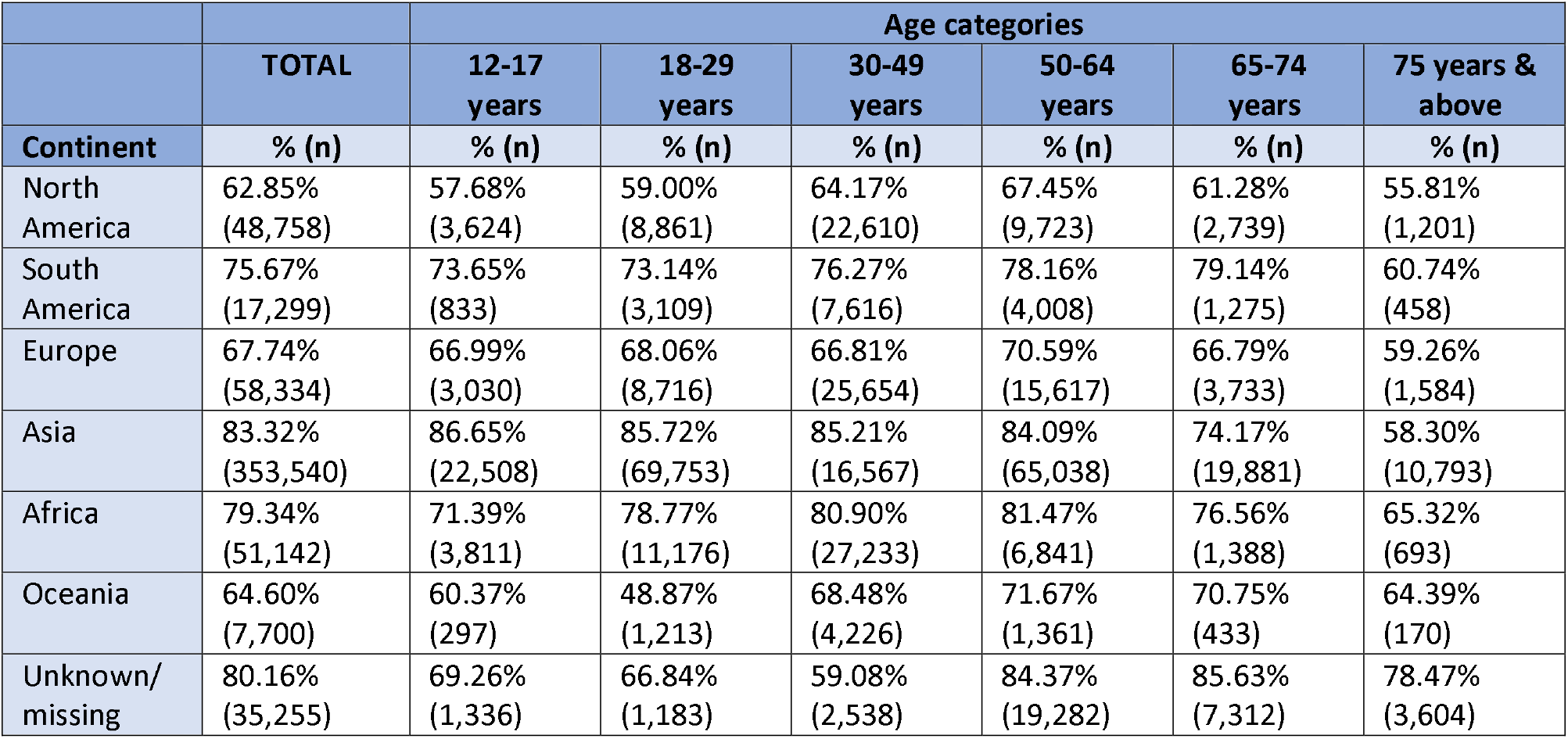
Vaccination coverage of international immigrants to Alberta categorized by continent of origin and age.

The proportions of vaccinated individuals were significantly different (p<0.001) by age and migration status (Table 2). Vaccination coverage for immigrants generally trended downwards after 50 years of age, irrespective of time since migration. Among immigrants under 50 years of age, those who migrated within the past 20 years tended to have higher vaccination coverage than those who migrated more than 20 years ago. However, among older immigrants (50 years of age and older), those who migrated within the past 20 years had lower coverage than those who migrated more than 20 years ago. In non-immigrants, vaccination coverage increased substantially in older age categories; 70.9%-73.5%% of those 12-49 years old were vaccinated, increasing thereafter, with 86.3% of those 75 years and above having received at least one dose.

**Table 2.** Proportion of Alberta residents who received at least one dose of a COVID-19 vaccine by migration status and sociodemographic characteristics.

|  | Immigrant categories by time since migration<br>(N = 731,217) |  |  |  | Non-immigrants<br>(N=3,200,481) | p-value |
| --- | --- | --- | --- | --- | --- | --- |
|  | <2<br>years | 3-10 years | 11-20 years | > 20 years |  |  |
| Variables | % (n) | % (n) | % (n) | % (n) | % (n) |  |
| **Age** |  |  |  |  |  |  |
| **12-17 years** | 81.99%<br>(3,546) | 79.80%<br>(23,422) | 70.63%<br>(8,471) | *N/A | 72.07%<br>(177,414) | <0.001 |
| **18-29 years** | 78.23%<br>(16,359) | 81.40%<br>(53,102) | 76.01%<br>(30,423) | 72.14%<br>(4,127) | 73.46%<br>(384,093) | <0.001 |
| **30-49 years** | 82.20%<br>(22,355) | 80.37%<br>(119,317) | 77.74%<br>(91,172) | 77.62%<br>(22,600) | 70.94%<br>(783,846) | <0.001 |
| **50-64 years** | 70.81%<br>(3,008) | 76.54%<br>(22,500) | 78.52%<br>(46,292) | 84.07%<br>(50,070) | 79.13%<br>(564,189) | <0.001 |
| **65-74 years** | 61.76%<br>(1,702) | 63.50%<br>(7,903) | 72.75%<br>(8,403) | 82.66%<br>(18,753) | 84.19%<br>(307,192) | <0.001 |
| **75 years & above** | 56.19%<br>(558) | 49.74%<br>(3,421) | 55.91%<br>(4,189) | 70.56%<br>(10,335) | 86.31%<br>(214,558) | <0.001 |
| **Sex** |  |  |  |  |  |  |
| Female | 78.63%<br>(24,105) | 79.27%<br>(120,856) | 77.23%<br>(97,798) | 81.52%<br>(56,494) | 77.75%<br>(1,223,034) | <0.001 |
| Male | 78.67%<br>(23,423) | 78.11%<br>(108,809) | 75.54%<br>(91,152) | 79.11%<br>(49,391) | 74.24%<br>(1,208,258) | <0.001 |
| **Place of residence** |  |  |  |  |  |  |
| Rural | 64.24%<br>(4,225) | 71.08%<br>(18,082) | 59.50%<br>(12,615) | 66.97%<br>(7,941) | 68.14%<br>(443,505) | <0.001 |
| Urban | 76.43%<br>(3,738) | 79.25%<br>(19,180) | 75.38%<br>(13,576) | 75.44%<br>(5,883) | 71.19%<br>(301,335) | <0.001 |
| Metro and moderate Metro | 80.81%<br>(39,565) | 79.46%<br>(192,403) | 78.22%<br>(162,759) | 82.14%<br>(92,061) | 79.31%<br>(1,686,452) | <0.001 |
| **Income quintile** |  |  |  |  |  |  |
| Q1 (lowest) | 80.15%<br>(13,625) | 79.35%<br>(61,772) | 74.88%<br>(41,535) | 79.97%<br>(22,785) | 71.43%<br>(382,968) | <0.001 |
| Q2 | 78.11%<br>(11,078) | 79.64%<br>(50,469) | 75.59%<br>(37,562) | 80.38%<br>(23,281) | 74.21%<br>(467,438) | <0.001 |
| Q3 | 78.23%<br>(7,983) | 78.62%<br>(40,920) | 76.52%<br>(36,741) | 80.26%<br>(21,730) | 76.38%<br>(499,438) | <0.001 |
| Q4 | 78.04%<br>(7,854) | 78.26%<br>(39,871) | 77.60%<br>(37,558) | 80.77%<br>(20,500) | 77.64%<br>(535,676) | <0.001 |
| Q5 (highest) | 77.83%<br>(6,988) | 77.03%<br>(36,633) | 77.77%<br>(35,554) | 80.61%<br>(17,589) | 79.03%<br>(545,772) | <0.001 |

\*There was no one in this age category among immigrants who migrated more than 20 years ago.

There was also variability by other sociodemographic characteristics and migration status. Vaccination coverage among female immigrants was slightly higher than males (except for those who migrated in the last 2 years), with coverage difference increasing slightly with increasing time since migration (Table 2). This difference between females and males was more pronounced in non-immigrants (77.8% vs 74.2%). By place of residence, metro areas had the highest vaccination coverage compared to urban and rural areas, regardless of migration status. There was a wider gap in immigrants who lived in rural areas compared to immigrants who lived in urban areas (76.4% vs 64.2% among those migrating in the last 2 years). Among non-immigrants, the gap between urban and rural residents was 3% (68.1% vs 71.2%). In immigrants, coverage was consistent across the income quintiles, while in non-immigrants, coverage ranged from 71.4% in the poorest quintile to 79.0% in the richest quintile.

### Interaction between age and migration status

There was a significant interaction between age category and migration status (Table 3 and Figure 1). Among immigrants, younger age groups that immigrated most recently (10 years or less) had higher coverage than those who immigrated more than 10 years ago. This trend changes after 50 years of age, when coverage in more recent immigrants starts to drop below those who immigrated more than 10 years ago. Among non-immigrants, vaccine coverage continues to increase with age, peaking in those 75 years and above (86.3%). Multivariable logistic regression analysis showed that immigrants generally had significantly higher vaccine uptake for those aged 12 to 49 years old compared to non-immigrants. However, among those aged 50 years and above, immigrants had significantly lower vaccine uptake compared to non-immigrants. Immigrants who migrated in the last 2 years had nearly twice the likelihood of getting vaccinated compared to non-immigrants (OR = 1.73, 95% CI: 1.61, 1.86) among those aged 12-17 years. Conversely, among those aged 75 years and above, immigrants who migrated in the last 2 years had an 83% lower chance of getting vaccinated compared to non-immigrants (OR = 0.17, 95% CI: 0.14, 0.19).

**Table 3.** Bivariate and multivariable association of vaccination with migration status.

| Migration status |  | Unadjusted OR | 95% CI | Adjusted OR* | 95% CI |
| --- | --- | --- | --- | --- | --- |
| Migrated in last 2 years | 12-17 years | 1.76 | 1.63-1.91 | 1.73 | 1.61-1.86 |
| Migrated in last 3-10 |  | 1.53 | 1.49-1.58 | 1.49 | 1.45-1.53 |
| Migrated in last 11-20 years |  | 0.93 | 0.90-0.97 | 0.90 | 0.87-0.93 |
| Non-migrants |  | Ref |  | Ref |  |
| Migrated in last 2 years | 18-29 years | 1.3 | 1.26-1.34 | 1.34 | 1.29-1.38 |
| Migrated in last 3-10 |  | 1.58 | 1.55-1.63 | 1.51 | 1.48-1.54 |
| Migrated in last 11-20 years |  | 1.15 | 1.12-1.17 | 1.07 | 1.04-1.09 |
| Migrated before 20 years |  | 0.94 | 0.88-0.99 | 0.88 | 0.83-0.93 |
| Non-migrants |  | Ref |  | Ref |  |
| Migrated in last 2 years | 30-49 years | 1.89 | 1.83-1.95 | 1.83 | 1.77-1.89 |
| Migrated in last 3-10 |  | 1.68 | 1.66-1.70 | 1.57 | 1.55-1.60 |
| Migrated in last 11-20 years |  | 1.43 | 1.41-1.45 | 1.34 | 1.32-1.36 |
| Migrated before 20 years |  | 1.42 | 1.38-1.46 | 1.34 | 1.31-1.38 |
| Non-migrants |  | Ref |  | Ref |  |
| Migrated in last 2 years | 50-64 years | 0.64 | 0.60-0.68 | 0.51 | 0.48-0.54 |
| Migrated in last 3-10 |  | 0.86 | 0.84-0.88 | 0.73 | 0.71-0.75 |
| Migrated in last 11-20 years |  | 0.96 | 0.94-0.98 | 0.83 | 0.81-0.84 |
| Migrated before 20 years |  | 1.39 | 1.36-1.42 | 1.24 | 1.21-1.27 |
| Non-migrants |  | Ref |  | Ref |  |
| Migrated in last 2 years | 65-74 years | 0.3 | 0.28-0.33 | 0.23 | 0.21-0.25 |
| Migrated in last 3-10 |  | 0.33 | 0.32-0.34 | 0.25 | 0.24-0.26 |
| Migrated in last 11-20 years |  | 0.5 | 0.48-0.52 | 0.42 | 0.40-0.43 |
| Migrated before 20 years |  | 0.9 | 0.86-0.93 | 0.77 | 0.74-0.80 |
| Non-migrants |  | Ref |  | Ref |  |
| Migrated in last 2 years | 75 years and above | 0.2 | 0.18-0.23 | 0.17 | 0.14-0.19 |
| Migrated in last 3-10 |  | 0.16 | 0.15-0.17 | 0.12 | 0.11-0.13 |
| Migrated in last 11-20 years |  | 0.2 | 0.19-0.21 | 0.16 | 0.15-0.17 |
| Migrated before 20 years |  | 0.38 | 0.37-0.39 | 0.31 | 0.30-0.33 |
| Non-migrants |  | Ref |  | Ref |  |

\*Adjusted for age category, sex, place of residence, and neighborhood income quintile.

**Figure 1.**
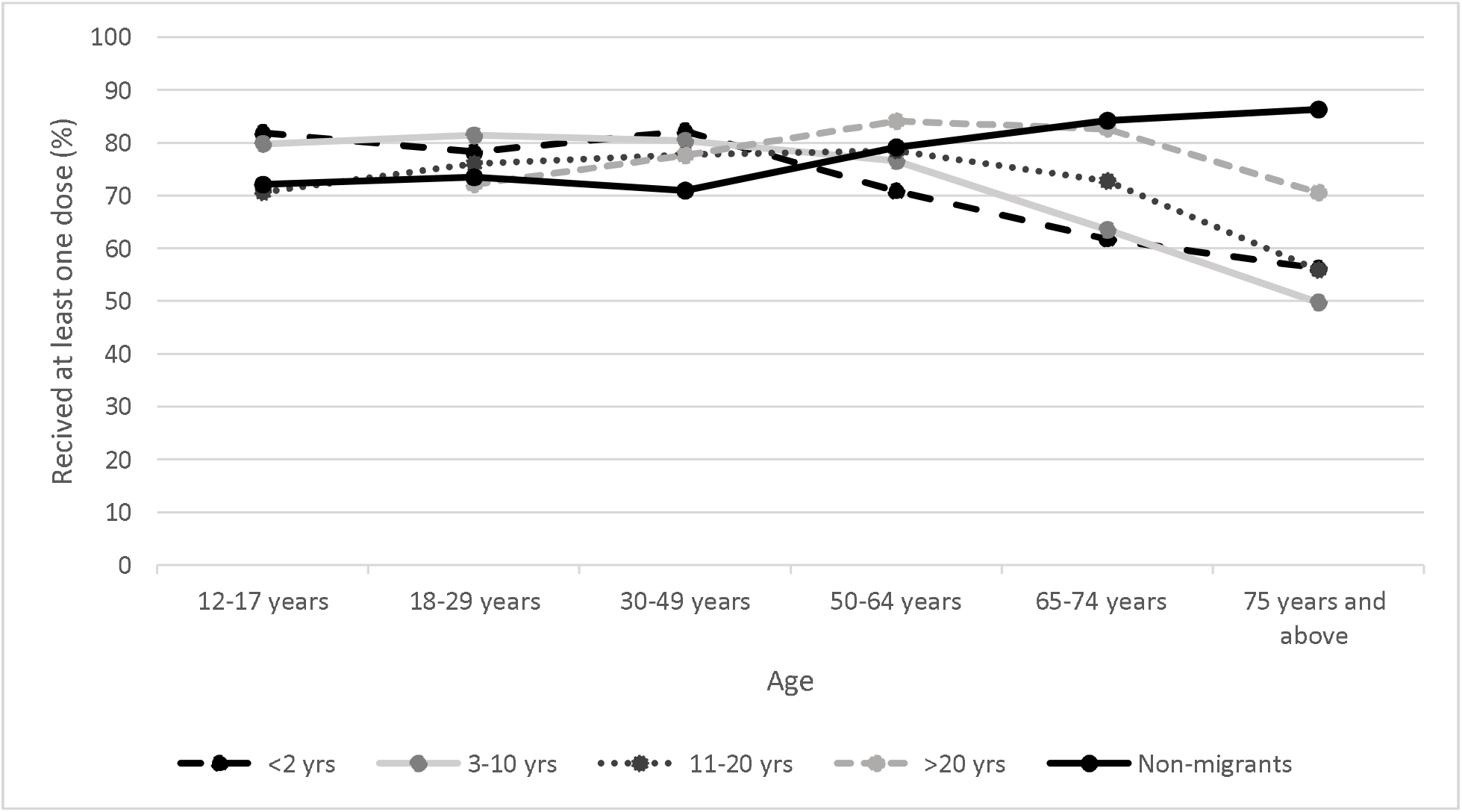
Vaccination coverage in different age categories by migration status.

## Discussion

### Summary of findings

This study reports on the status of COVID-19 vaccination coverage among immigrants in comparison to non-immigrants in Alberta. Overall, vaccine coverage among immigrant populations is slightly higher compared to non-immigrants and was significantly higher among immigrants under 50 years of age. Unlike the non-immigrant population, which had very high coverage in older adults, coverage decreased in older immigrants, regardless of time since migration.

### Interpretation

Previous studies have shown lower uptake of routine vaccines among immigrant populations compared to non-immigrants.(6,12–14) However, COVID-19 vaccination intention in a study from California, USA was higher among immigrant adults (75%) compared to non-immigrant adults (68%).(15) Similarly, a Canadian web survey of participants aged 18 years and older also found that immigrants had a higher intention to receive the COVID vaccine (75%) compared to Canadian-born residents (69%). The difference in intention was the highest in Alberta (86% in immigrants versus 61% in Canadian-born).(16) The same survey reported that nearly 80% of the immigrants were satisfied with the federal government’s handling of the pandemic.(17) Immigrants in Alberta generally have a high trust towards government and health authorities regarding vaccination decisions.(18) Additionally, immigrants are more likely to be employed in essential occupations where there are higher chances of exposure to COVID-19 infection,(19) which could have motivated immigrants to get vaccinated.(20) Some employers instituted vaccine mandates for workers in essential occupations at the beginning of the pandemic. As well, the availability of vaccines free-of-charge through public health centers, pharmacies, and physician clinics irrespective of migration status might have reduced barriers. Although studies have not further explored COVID-19 vaccine hesitancy between immigrants and non-immigrants, differing concerns about safety, efficacy, and perception of mild disease may also explain the differences.(21)

Immigrants from Europe, Oceania, and North America had lower vaccine coverage for those aged 12 to 74 years, compared to immigrants from Asia. Older immigrants (75 years and above) had lower vaccine coverage than middle-aged (30-49 years) immigrants for all continents of origin. Consistent with our findings of higher overall coverage in Asian immigrants, an earlier study from Norway found that immigrants from Asian countries (Vietnam, Sri Lanka, Thailand) had COVID-19 vaccine coverage similar to Norwegian-born individuals (>90%).(22) They also found that vaccine coverage among immigrants from other Asian countries (India, Philippines, Iran) and from Northern Europe (Denmark, UK, Sweden) was much higher than immigrants from Eastern Europe (Latvia, Bulgaria, Poland, Romania, and Lithuania, and proposed that vaccine hesitancy translated into lower vaccine uptake.(22) Vaccine hesitancy in these countries is believed to be associated with a history of authoritarian governments.(23,24)

Older immigrants (65 years and older) had significantly lower coverage compared to older non-immigrants and this difference was higher among recent immigrants. This finding is particularly relevant as older age groups were one of the first priority groups for COVID-19 vaccination in Alberta. A study from Ontario, Canada also showed that COVID-19 vaccine coverage was significantly lower among immigrants and refugees aged 70 years and over, compared to their Canadian-born counterparts.(25) Lower uptake of pneumococcal vaccine among older immigrants was also reported in the US.(14) Possible reasons for lower uptake among older immigrants may be related to cultural and language barriers, higher exposure to misinformation from informal sources and social media, distance to clinics/pharmacies, opening hours, and challenges with online booking systems.(26) Additionally, residents in long term care (LTC) and designated supported living (DSL) facilities were the highest priority groups for COVID-19 vaccination in Canada.(27) Lower admission rate among older immigrants in LTC and DSL facilities due to their ‘strong preference to grow old at home’ and LTC-related barriers might have resulted in lower coverage.(28)

We found that vaccination coverage by migration status showed an interaction with age category. Vaccination coverage among immigrants who migrated in the last 10 years started to drop after 50 years of age. Among non-immigrants, vaccine coverage rose after 50 years of age. Interestingly, immigrants who migrated more than 20 years ago followed a pattern similar to non-immigrants, i.e., increasing coverage after 50 years of age, until the oldest age groups, when coverage started to decline. Since views and beliefs about vaccines are influenced by interpersonal relationships, as well as an individual’s historical and social context,(29) it is possible that migrating later in life means that early lifetime influences of older immigrants continue to affect their vaccination behaviour to present day. Among younger immigrants, however, higher educational status before migration, and smooth acclimatization and integration into Canadian society may have instilled positive views towards vaccines. It can thus be suggested that factors such as time since migration, age at migration, and acculturation window determine the occurrence of the healthy immigrant effect in terms of COVID-19 vaccination, and it does not hold for older immigrants.

### Future directions

Understanding how COVID-19 vaccine uptake varies in Canada’s immigrant population is important to design strategies for vaccine delivery and messaging.(12) This is particularly true for older immigrants who had disturbingly low coverage levels, despite their high risk from COVID-19 infection. Our findings suggest that COVID-19 vaccination strategies need to be tailored to the needs of diverse immigrant populations. This could include translating educational materials about vaccines into the language they understand.(16) Strategies may vary for immigrants from North America, Oceania, and Europe, who may not face language barriers, but nonetheless had lower coverage. Moreover, rural-residing immigrants might need additional strategies to receive the COVID-19 vaccine. These might include co-designing tailored approaches according to local needs, leveraging the role of social media, reconsidering current models for vaccine distribution,(30) and partnering with local community-based organizations,(26) faith leaders, and underserved communities, including immigrant communities either directly or through local partners.(31) Communication through text messages, mass media, or telephone, as well as offering vaccines on a walk-in basis can attract diverse and hard-to-reach populations.(32)

### Strengths and limitations

This is one of the few studies in Canada to investigate vaccine coverage across different age groups of immigrant populations in comparison to non-immigrant populations. We used population-based immunization, immigrant, and resident registries to derive our coverage estimates, ensuring a large and complete dataset for analysis.

Our study has some limitations. Our dataset likely included individuals who moved out of Alberta without reporting this to the provincial health insurance registry, which may have resulted in an underestimation of vaccine coverage in all groups. Thus, our denominator is larger than reported by Alberta Health,(33) which applies aggregate weights to offset the population inflation, an approach that was not possible for our individual-level data. We did attempt to remove non-residents from our denominator through other approaches. For instance, we tried excluding individuals aged 50 years and over who had not used any of the health services in seven domains in 2020 and 2021 (ambulatory care, inpatient care, laboratory data, COVID-19 test data, long term care, pharmacy data, physician claims), assuming that if they did not use any of these health services then they were probably living out of the province. However, although vaccine coverage by immigration status showed a similar pattern as was seen in the full sample, we lost a significant number of individuals who received COVID-19 vaccines in Alberta in the past year. Therefore, we decided to conduct analysis on the full sample.

In addition, since interprovincial migration was included in the non-immigrant population, there may have been some misclassification of international immigrants as non-immigrants, thereby overestimating the non-immigrant coverage levels. Furthermore, since children born to immigrants in Canada are included in the non-immigrant populations, it might have also overestimated the vaccine coverage among non-immigrants. The findings from this study may not be generalizable to other jurisdictions due to differences in immigration trends, and immigrant characteristics, among others. Finally, COVID-19 vaccines received outside of Alberta may not have been captured in Alberta’s immunization repository.

### Conclusion

Public health interventions should focus on older immigrants, immigrants living in rural areas, and immigrants from specific continental backgrounds to improve COVID-19 vaccination coverage. Future research should examine COVID-19 vaccination uptake in children of immigrants, specifically the newly eligible 5- to 11-year-old age group.

## Data Availability

Data may be available from the Alberta Ministry of Health upon request and are not publicly available.

## Acknowledgements

We would like to acknowledge Dr. Larry Svenson for his role in conceptualizing and initiating this project. It was one of the many interesting project ideas that he developed in his career at Alberta Health. We are mourning his recent death for many reasons, not the least of which is the loss of his creative mind. We will miss the man we called “the idea guy”.

## Contributors

All authors attest that they meet the ICMJE criteria for authorship.

SEM was involved in conceptualization, methodology, investigation, funding acquisition, writing (review and editing), supervision.

YRP was involved in conceptualization, methodology, investigation, formal analysis, writing (original draft, review, and editing).

CD assisted with data interpretation and writing (original draft, review, and editing).

## Ethics statement

This research, involving data from human participants, conformed to the principles embodied in the Declaration of Helsinki and was granted ethical approval by the University of Alberta Health Research Ethics Board (Ethics ID: Pro00114786).

## References

1. Bolotin S, Wilson S, Murti M. Achieving and sustaining herd immunity to SARS-CoV-2. CMAJ. 2021;193(28):E1089. doi: 10.1503/cmaj.210892

2. Statistics Canada. Immigration and ethnocultural diversity: Key results from the 2016 Census [Internet]. 2017 Oct [cited 2022 Mar 14]. Available from: https://www.statcan.gc.ca/en/subjects-start/immigration_and_ethnocultural_diversity

3. Chief Public Health Officer. From risk to resilience: An equity approach to COVID-19 [Internet]. Public Health Agency of Canada; 2020 [cited 2022 Mar 14]. p. 1–96. Available from: https://www.canada.ca/en/public-health/corporate/publications/chief-public-health-officer-reports-state-public-health-canada/from-risk-resilience-equity-approach-covid-19.html

4. Ng E, Sanmartin C, Elien-Massenat D, Manuel DG. Vaccine-preventable disease-related hospitalization among immigrants and refugees to Canada: Study of linked population-based databases. Vaccine. 2016;34(37):4437–42.

5. Greenaway C, Greenwald ZR, Akaberi A, Song S, Passos-Castilho AM, Abou Chakra CN, et al. Epidemiology of varicella among immigrants and non-immigrants in Quebec, Canada, before and after the introduction of childhood varicella vaccination: a retrospective cohort study. Lancet Infect Dis. 2021;21(1):116–26.

6. Thomas CM, Osterholm MT, Stauffer WM. Perspective piece critical considerations for COVID-19 vaccination of refugees, immigrants, and migrants. Am J Trop Med Hyg. 2021;104(2):433–5. doi: 10.4269/ajtmh.20-1614

7. Wilson L, Rubens-Augustson T, Murphy M, Jardine C, Crowcroft N, Hui C, et al. Barriers to immunization among newcomers: A systematic review. Vaccine. 2018;36(8):1055–62.

8. Government of Alberta. A COVID-19 vaccine: Phase 2B and 2C [Internet]. Government of Alberta; 2020 [cited 2022 Mar 14]. Available from: https://abrda.ca/wp-content/uploads/2021/03/Albertas-COVID-19-Vaccine-Phase-2B-and-2C-Information.pdf

9. Government of Alberta. COVID-19 Alberta statistics [Internet]. Available from: https://www.alberta.ca/stats/covid-19-alberta-statistics.htm#vaccinations

10. Detsky AS, Bogoch II. COVID-19 in Canada: Experience and response to waves 2 and 3. JAMA. 2021;326(12):1145–6. doi: 10.1001/jama.2021.14797

11. Adeyinka DA, Camillo CA, Marks W, Muhajarine N. Implications of COVID-19 vaccination and public health countermeasures on SARS-CoV-2 variants of concern in Canada: evidence from a spatial hierarchical cluster analysis [Preprint]. medRxiv. Available from: https://doi.org/10.1101/2021.06.28.21259629

12. Karki S, Dyda A, Newall A, Heywood A, MacIntyre CR, McIntyre P, et al. Comparison of influenza vaccination coverage between immigrant and Australian-born adults. Vaccine. 2016;34(50):6388–95.

13. Mereckiene J, Cotter S, D’Ancona F, Giambi C, Nicoll A, Levy-Bruhl D, et al. Differences in national influenza vaccination policies across the European Union, Norway and Iceland 2008-2009. Euro Surveill. 2010;15(44):19700. doi: 10.2807/ese.15.44.19700-en

14. Lu PJ, Rodriguez-Lainz A, O’Halloran A, Greby S, Williams WW. Adult vaccination disparities among foreign-born populations in the U.S., 2012. Am J Prev Med. 2014;47(6):722–33.

15. Gonzalez D, Karpman M, Bernstein H. COVID-19 vaccine attitudes among adults in immigrant families in California [Internet]. Urban Institute; 2021 [cited 2022 Mar 14]. Available from: https://www.urban.org/sites/default/files/publication/103973/covid-19-vaccine-attitudes-among-adults-in-immigrant-families-in-california_0_0.pdf

16. Viseman N. Vaccine intentions among Canadian immigrants in COVID-19: Why does it matter? [Internet]. University of Manitoba; 2021 [cited 2022 Mar 14]. Available from: https://news.umanitoba.ca/wp-content/uploads/2020/10/2020-UMUPC-SSH-VEISMAN-NIKOL.pdf

17. Griffith J, Marani H, Monkman H. COVID-19 vaccine hesitancy in Canada: Content analysis of tweets using the theoretical domains framework. J Med Internet Res. 2021;23(4):e26874. doi: 10.2196/26874.

18. Kowal SP, Jardine CG, Bubela TM. “If they tell me to get it, I’ll get it. If they don’t….”: Immunization decision-making processes of immigrant mothers. Can J Public Health. 2015;106(4):e230–5.

19. Hayward SE, Deal A, Cheng C, Crawshaw A, Orcutt M, Vandrevala TF, et al. Clinical outcomes and risk factors for COVID-19 among migrant populations in high-income countries: A systematic review. J Migr Health. 2021;3:100041.

20. Brewer NT, Chapman GB, Gibbons FX, Gerrard M, McCaul KD, Weinstein ND. Meta-analysis of the relationship between risk perception and health behavior: The example of vaccination. Health Psychol. 2007;26(2):136–45.

21. Dror AA, Eisenbach N, Taiber S, Morozov NG, Mizrachi M, Zigron A, et al. Vaccine hesitancy: The next challenge in the fight against COVID-19. Eur J Epidemiol. 2020;35(8):775–9. doi: 10.1007/s10654-020-00671-y

22. Kraft B, Godøy A, Vinjerui K, Kour P, Kjøllesdal M, Indseth T. COVID-19 vaccination coverage by immigrant background. Tidsskr Nor Laegeforen. 2021;141(2). doi: 10.4045/tidsskr.21.0799

23. Kroeger A. Vaccine hesitancy in south-eastern Europe stems from the region’s authoritarian past [Internet]. 2021 [cited 2022 Apr 8]. Available from: https://www.newstatesman.com/world/europe/2021/11/low

24. Andjelic N. Democracy, Liberties and Rights Under the Threat of State Fighting the Pandemic. Vol. 5. Cham Springer; 2022. 137–163 p.

25. Institute for Clinical Evaluative Sciences. Vaccine Coverage by Neighbourhood COVID-19 Risk in Immigrants, Refugees, and other Newcomers, up to April 26, 2021 [Internet]. 2021 [cited 2022 Apr 8]. Available from: https://www.ices.on.ca

26. Crawshaw AF, Deal A, Rustage K, Forster AS, Campos-Matos I, Vandrevala T, et al. What must be done to tackle vaccine hesitancy and barriers to COVID-19 vaccination in migrants? J Travel Med. 2021;28(4):taab048. doi: 10.1093/jtm/taab048

27. Lauter S, Lorenz-Dant K, Comas-Herrera A, Perobelli Eleonora, Caress A, Sinha S, et al. International “living” report: Long-term care and COVID-19 vaccination, prioritization and data, 26th January 2021 update [Internet]. International Long-Term Care Policy Network; 2021 [cited 2022 Mar 23]. Available from: https://ltccovid.org/2021/01/26/new-international-living-report-long-term-care-and-covid-19-vaccination-prioritization-and-data/

28. Salma J, Salami B, Bowers BJ. “We are like any other people, but we don’t cry much because nobody listens”: The need to strengthen aging policies and service provision for minorities in Canada. Gerontologist. 2020;60(2):279–90.

29. Dubé E, Laberge C, Guay M, Bramadat P, Roy R, Bettinger JA. Vaccine hesitancy: An overview. Hum Vaccin Immunother. 2013;9(8):1763–73. doi: 10.4161/hv.24657

30. Prusaczyk B. Strategies for disseminating and implementing COVID-19 vaccines in rural areas. Open Forum Infect Dis. 2021;8(6):ofab152. doi: 10.1093/ofid/ofab152

31. Patel Murthy B, Sterrett N, Weller D, Zell E, Reynolds L, Toblin RL, et al. Disparities in COVID-19 vaccination coverage between urban and rural counties — United States, December 14, 2020– April 10, 2021. MMWR. 2021;70(20):759–64.

32. Stern RJ, Rafferty HF, Robert AC, Taniguchi C, Gregory B, Khoong EC, et al. Concentrating vaccines in neighborhoods with high Covid-19 burden [Internet]. NEJM Catalyst. 2021. Available from: https://catalyst.nejm.org/doi/full/10.1056/CAT.21.0056

33. Government of Alberta. Interactive Health Data Application [Internet]. Government of Alberta; 2021 [cited 2022 Mar 15]. Available from: http://www.ahw.gov.ab.ca/IHDA_Retrieval/selectCategory.do

