## Supplemental tables and figures for "COVID-19 vaccine coverage among immigrants and refugees in Alberta: a population-based cross-sectional study"

**Appendix**

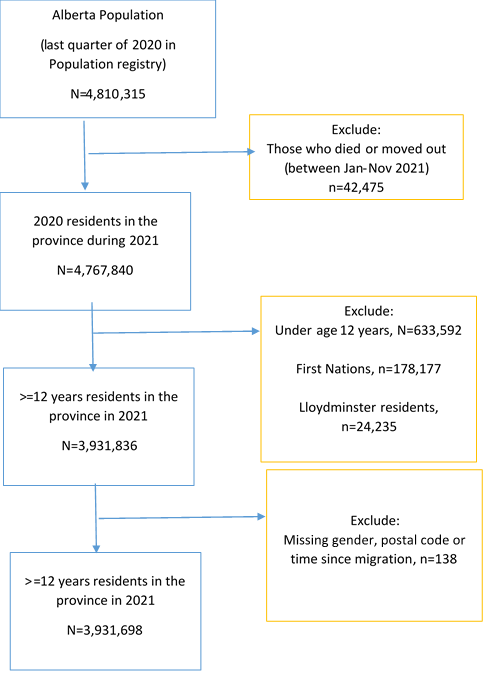

**Figure A1. Flow diagram showing selection of participants.**

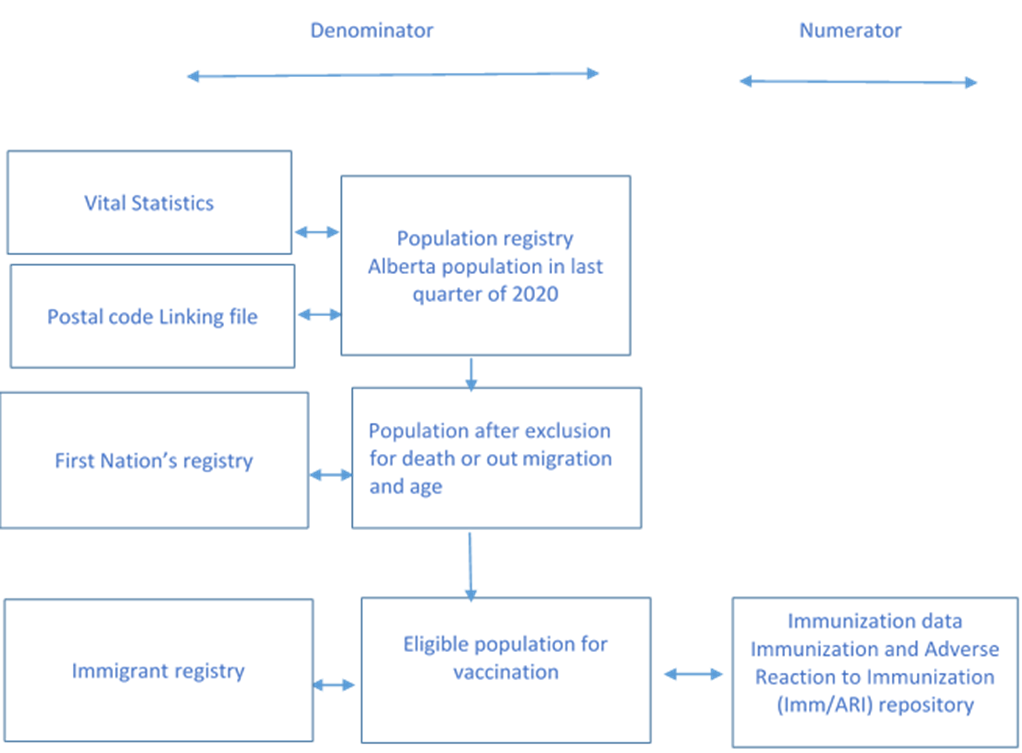

**Figure A2. Linkage of different databases.**

**Table A1: Vaccination coverage of Albertan immigrants categorized by sub-continental region of origin**

| **Continent** | **% (n)** |
| --- | --- |
| North America | 63.19% (43,439) |
| Meso America | 62.42% (34,137) |
| South America | 75.67% (17,299) |
| Europe Unspecified | 72.45% (4,191) |
| East Europe | 60.69% (11,512) |
| West Europe | 64.05% (9,417) |
| North Europe | 71.50% (24,356) |
| South Europe | 70.34% (8,858) |
| Central Asia | 64.59% (5,370) |
| Middle East | 75.67% (41,263) |
| Africa Unspecified | 79.70% (2,308) |
| North Africa | 75.41% (7,825) |
| East Africa | 79.51% (20,122) |
| Central/South Africa | 76.41% (6,775) |
| West Africa | 82.94% (14,112) |
| Asia Unspecified | 86.21% (11,857) |
| East Asia | 85.47% (193,589) |
| South Asia | 83.71% (101,461) |
| Oceania | 64.60% (7,700) |
| Unknown/Missing | 80.16% (35,255) |
